# Ultra-processed food staples dominate mainstream U.S. supermarkets. Americans more than Europeans forced to choose between health and cost

**DOI:** 10.1101/2024.02.16.24302894

**Authors:** Bertrand Amaraggi, Wendy Wood, Laura Guinovart Martín, Jaime Giménez Sánchez, Yolanda Fleta Sánchez, Andrea de la Garza Puentes

## Abstract

**BACKGROUND:** The United States (U.S.) is the leading country in ultra-processed food (UPF) consumption, accounting for 60% of caloric intake, compared to a range of 14 to 44% in Europe. Given the increasingly evident health risks of UPF consumption, this is a major health problem. Common UPFs include soft drinks, snacks, processed meats, cookies, and candy. We hypothesized that even basic staple foods in the United States are ultra-processed and aimed to study the UPF prevalence in food staples from popular U.S. supermarkets compared with European countries.

**METHODOLOGY:** We analyzed staple food products (bread, canned goods, cereals, eggs, milk, vegetables, and yogurt) stocked in U.S. supermarket chains (Walmart, Target, and Whole Foods), France (Carrefour), and Spain (Mercadona). Using an algorithm that identifies UPF values based on the NOVA and Food and Agriculture Organization of the United Nations guidelines, we identified the UPF prevalence and average number of UPF markers (cosmetic ingredients/additives).

**RESULTS:** The prevalence of UPFs in budget-friendly supermarkets, Walmart, and Target, is 41-42% higher than Whole Foods, a store focused on quality. Furthermore, UPFs in Walmart and Target have 75% and 57% more UPF markers, respectively, than UPFs in Whole Foods. Around 58% of staples in U.S. leading supermarkets are ultra-processed, which is 41% more than supermarkets in Europe. Furthermore, the U.S. UPFs contain 41% more UPF markers than their EU counterparts.

**CONCLUSION:** Most of the staple food products at mainstream U.S. budget-friendly retailers are ultra-processed, which is not the case at a more premium, quality focused store. Compared to supermarkets in Europe, the U.S. mainstream supermarkets have more UPFs, and those foods also have more UPF markers. Making healthy food choices in the United States is a challenge that is compromised by the high availability and accessibility of UPFs, even among everyday products that constitute the dominant part of the diet of a population. The European model shows the possibility of decreasing the UPF availability in large supermarkets. American consumers need more tools and guidance to identify UPFs along with greater regulation of UPF products to prioritize healthy choices and reduce UPF availability.

## 1 INTRODUCTION

The United States (U.S.) is the leading country in ultra-processed food (UPF) consumption (1), with UPFs accounting for about 60% of total daily caloric intake. These numbers are even higher among consumers with lower income and education (2). In Europe, countries have a lower UPF consumption ranging from 14 to 44% of energy share (3). These rates are a major health problem given the increasing evidence of a link between the intake of UPFs and adverse health outcomes (4,5).

According to the Food and Agriculture Organization of the United Nations (FAO), UPFs are formulations of ingredients, mostly of exclusive industrial use, typically created by a series of industrial techniques and processes, hence ‘ultra-processed’. UPFs are characterized by the presence of ultra-processed markers, such as extracts and natural aromas, synthetic aromas, glucose syrup, native starches, and dextrose (6). The ingredients of UPFs are either food substances of little culinary use or types of additives that make the products more sellable, palatable, or hyper-palatable. The processes and the ingredients used to make UPFs create products convenient and attractive for consumers (e.g., ready to eat products) and profitable for manufacturers (e.g., low-cost ingredients, long shelf-life), but they also typically make the UPFs nutritionally unbalanced (7).

The consumption of these products is associated with non-communicable diseases (NCDs) and conditions, such as a cardiometabolic risk profile, cardiovascular disease, cerebrovascular disease, obesity (8), type 2 diabetes (9), cancer (10), and all-cause mortality (5). Moreover, UPF consumption has been associated with increased mental health disorders, such as depression and anxiety (11). These adverse consequences are not only due to UPF’s negative effect per se, but also because of the displacement of other, healthier foods from the diet. For instance, research has found an inverse association between UPF consumption and specific nutrients, for example protein, vitamins, dietary fiber, minerals, and overall nutrient-balanced patterns (12,13). UPF consumption also leads to the impactful burden of the NCDs, which are related to 74% of deaths globally (14) and a $47T estimated loss in economic output by 2030 (15). The morbidity, mortality, and economic costs necessary to treat these NCDs continue to climb (16), due to both the loss of economic productivity and the increased healthcare expenditure. UPFs clearly contribute to this economic and social burden given their link to NCDs and other conditions.

While UPFs tend to be less satiating (17), they also have a significantly lower price than unprocessed foods (18), which can play a role in choices by the food industry and consumers. Moreover, although consumers might know UPFs to be unhealthy, most cannot easily identify them and understand their implications (19). The most common UPFs include soft drinks, sweet or salty packaged snacks, candies, and pre-prepared food (20). Given that these make up a substantial part of American diets (21), it is no surprise that the population consumes high levels of UPFs (22), but what about everyday products that are the foundation of a population’s diet, such as bread, canned food, milk, and eggs? Can Americans reduce their UPF consumption by sticking with these more basic foods? Is there a difference between big budget retailers and more expensive supermarkets focused on quality? What is the situation in other countries?

We anticipated a high prevalence of UPFs even among staple products in the United States, especially in the country’s largest retailers that reach a big proportion of the population by stressing budget shopping. Such a pattern could force citizens to choose between shopping for more affordable or healthier foods. Therefore, we aimed to analyze the availability of UPFs and the number of UPF markers in staple products from different supermarkets in the United States. Additionally, to evaluate whether a trade-off between health and food affordability is inevitable, we compared staple foods in the U.S. mainstream supermarkets with corresponding major supermarkets in Europe (France and Spain).

## 2 METHODOLOGY

### 2.1 Study design and inclusion criteria

The study analyzed the prevalence of UPFs in U.S. supermarkets as well as the number of UPF markers identified in the individual products to explore the degree of processing as suggested by other authors (6). We then compared the situation with European countries, such as France and Spain.

To understand the food availability in the United States, we selected two of the largest, mainstream, supermarket retail chains focused on affordable choices, such as Walmart (23–25) and Target (24,26), and a higher end supermarket focused on health and quality, Whole Foods (27–29).

For the comparison with Europe, we focused on mainstream budget-friendly supermarkets. We selected Carrefour (30) and Mercadona (31) in France and Spain, respectively, and Walmart and Target in the United States.

The analyses focused on staple food products. Given that the product classification may vary from shop to shop, we included staple food products present in all the selected supermarkets: Bread, Canned goods, Cereals, Eggs, Milk, Vegetables, and Yogurt. As part of the selection criteria, we excluded subcategories not found in all supermarkets. For instance, we removed the subcategory “Bars” from the category “Cereals”, and “Brioche” and “Pizza” from the “Bread” category. To be able to detect if a product was ultra-processed, we also excluded products with missing information (e.g., missing ingredients) or with obvious errors (e.g., name or nutritional table instead of ingredients in the ingredients field).

### 2.2 Data collection

We obtained the information from the publicly available data on the online websites of Walmart, Target, Whole Foods, Mercadona, and Carrefour. From a first total of 11 874 products identified, we eliminated those that did not have the necessary information to be rated and kept the sub-categories of products present in all supermarkets to compare equivalent products. The final sample was 9 601 products across 5 supermarkets: 3,114 in Walmart (U.S.), 1,604 in Target (U.S.), 1,545 in Whole Foods (U.S.), 2,709 in Carrefour (France), and 625 in Mercadona (Spain). All of these represented the same 7 staple categories of food.

### 2.3 Identification of UPFs and UPF markers

To classify products and identify UPFs, we used the algorithm integrated in the app, GoCoCo (32), which is based on the NOVA classification and FAO guidelines (Table 1). According to the FAO (20), products are ultra-processed if their ingredients contain at least one item characteristic of UPFs. Using this logic, our algorithm analyzed the ingredients and detected UPF markers (cosmetic ingredients and/or additives) inside each product to identify UPFs and explore the degree of processing. For the latter analysis, five UPF marker categories were created according to the number of UPF markers present in the individual products: UPFs with 1 UPF marker, UPFs with 2 UPF markers, UPFs with 3 UPF markers, UPFs ≥ 3 UPF markers, and UPFs with ≥ 4 UPF markers.

**Table 1.**
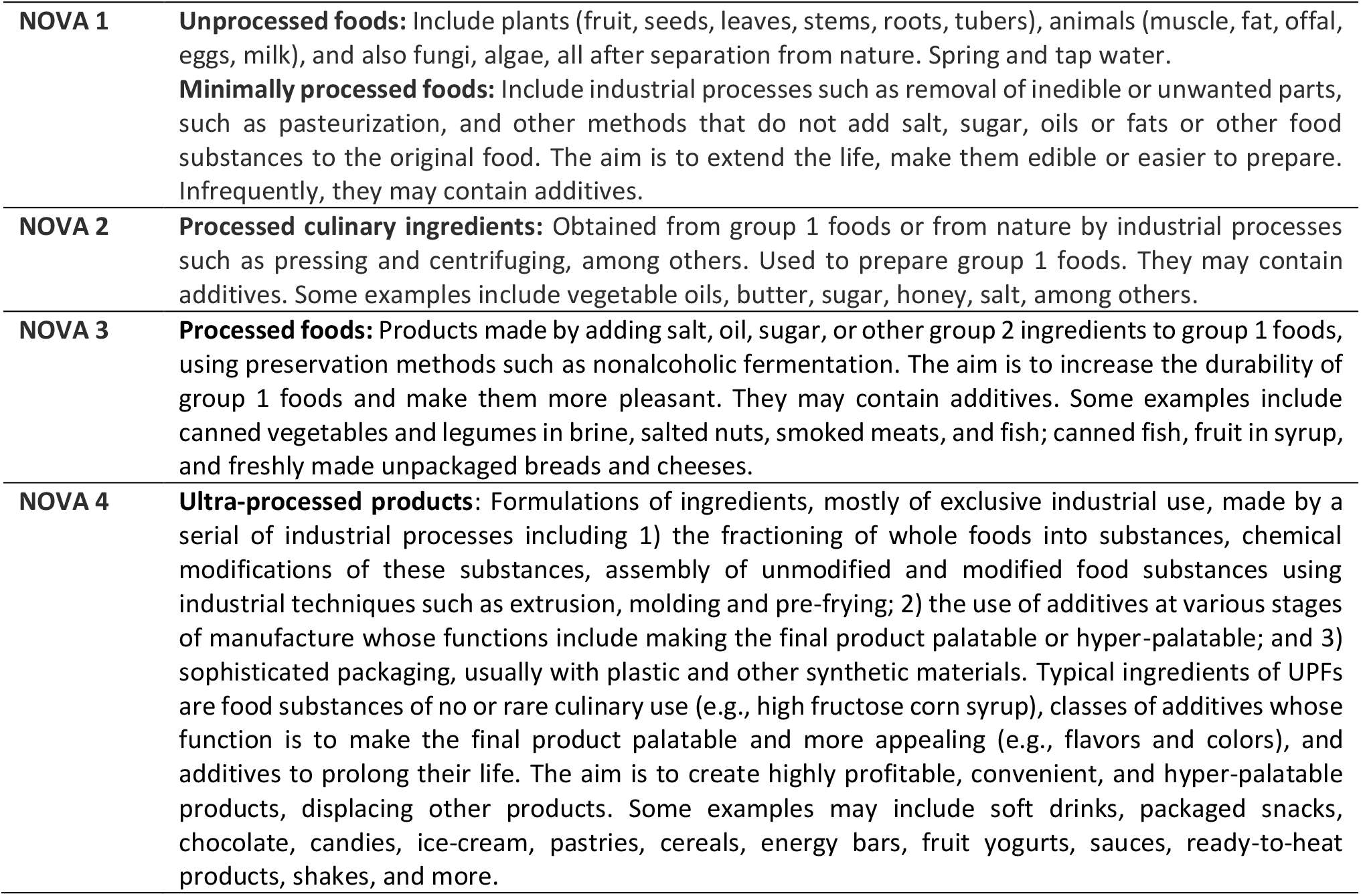
NOVA classification (7).

|  |  |
| --- | --- |
| **NOVA 1** | **Unprocessed foods:** Include plants (fruit, seeds, leaves, stems, roots, tubers), animals (muscle, fat, offal, eggs, milk), and also fungi, algae, all after separation from nature. Spring and tap water.<br>**Minimally processed foods:** Include industrial processes such as removal of inedible or unwanted parts, such as pasteurization, and other methods that do not add salt, sugar, oils or fats or other food substances to the original food. The aim is to extend the life, make them edible or easier to prepare. Infrequently, they may contain additives. |
| **NOVA 2** | **Processed culinary ingredients:** Obtained from group 1 foods or from nature by industrial processes such as pressing and centrifuging, among others. Used to prepare group 1 foods. They may contain additives. Some examples include vegetable oils, butter, sugar, honey, salt, among others. |
| **NOVA 3** | **Processed foods:** Products made by adding salt, oil, sugar, or other group 2 ingredients to group 1 foods, using preservation methods such as nonalcoholic fermentation. The aim is to increase the durability of group 1 foods and make them more pleasant. They may contain additives. Some examples include canned vegetables and legumes in brine, salted nuts, smoked meats, and fish; canned fish, fruit in syrup, and freshly made unpackaged breads and cheeses. |
| **NOVA 4** | **Ultra-processed products:** Formulations of ingredients, mostly of exclusive industrial use, made by a series of industrial processes including 1) the fractioning of whole foods into substances, chemical modifications of these substances, assembly of unmodified and modified food substances using industrial techniques such as extrusion, molding and pre-frying; 2) the use of additives at various stages of manufacture whose functions include making the final product palatable or hyper-palatable; and 3) sophisticated packaging, usually with plastic and other synthetic materials. Typical ingredients of UPFs are food substances of no or rare culinary use (e.g., high fructose corn syrup), classes of additives whose function is to make the final product palatable and more appealing (e.g., flavors and colors), and additives to prolong their life. The aim is to create highly profitable, convenient, and hyper-palatable products, displacing other products. Some examples may include soft drinks, packaged snacks, chocolate, candies, ice-cream, pastries, cereals, energy bars, fruit yogurts, sauces, ready-to-heat products, shakes, and more. |

### 2.4 Statistical analysis

The statistical analyses were performed using a custom script developed with Python programming language. For each supermarket in each country, we analyzed the distribution of products according to NOVA classification and the number of UPF markers. To examine the proportions of UPFs (classified as NOVA 4) we used the total amount of products and the count of UPFs in each supermarket and then performed *z*-tests to determine the differences between the selected supermarkets. This approach is particularly suitable for large sample sizes. Independent *t*-tests were also performed to compare the number of UPF markers present in the UPFs of each supermarket. For the analyses, we added a food category weight control to eliminate any potential differences due to the number of products in each category in the different supermarkets. The weighted analyses thus ensure that each category represents the same proportion in the different supermarkets, and in this way makes the values comparable. The significance level was established at *p* < 0.05, although most of the calculated *p* values were < 0.00001. This indicates that the differences were highly statistically significant and unlikely due to random chance.

## 3 RESULTS

### 3.1 UPF prevalence in the United States

Table 2 presents the distribution of the products included in the study. The prevalence of NOVA 4 products (UPFs) at Whole Foods ranged between 0 and 81% for individual food categories, yielding an overall average of 41%. In contrast, Walmart’s range was 3% to 92% and Target’s range was 0 to 98%, with overall averages in each store of 58%. Thus, in staple foods categories, Walmart had 42% more UPFs and Target 41% more than Whole Foods (see Table 3).

**Table 2.** Product distribution per NOVA category in the US supermarkets.

|  | Walmart<br>(n= 3114) | Target<br>(n= 1604) | Whole Foods<br>(n= 1545) |
| --- | --- | --- | --- |
| **NOVA 1** | 469 (15%) | 203 (13%) | 346 (22%) |
| **NOVA 3** | 824 (26%) | 468 (29%) | 561 (36%) |
| **NOVA 4** | **1 821 (58%)** | **933 (58%)** | **638 (41%)** |
| Bread | 190 (92%) | 112 (98%) | 112 (81%) |
| Canned goods | 479 (42%) | 278 (43%) | 109 (20%) |
| Cereals | 726 (74%) | 277 (73%) | 146 (59%) |
| Eggs | 2 (7%) | 0 (0%) | 0 (0%) |
| Milk | 46 (37%) | 25 (27%) | 20 (26%) |
| Vegetables | 8 (3%) | 12 (9%) | 0 (0%) |
| Yogurt | 370 (94%) | 229 (96%) | 251 (75%) |

The values presented are the number of products per category and the proportion they represent over the total amount. The food groups belong to the NOVA 4 category.

**Table 3.**
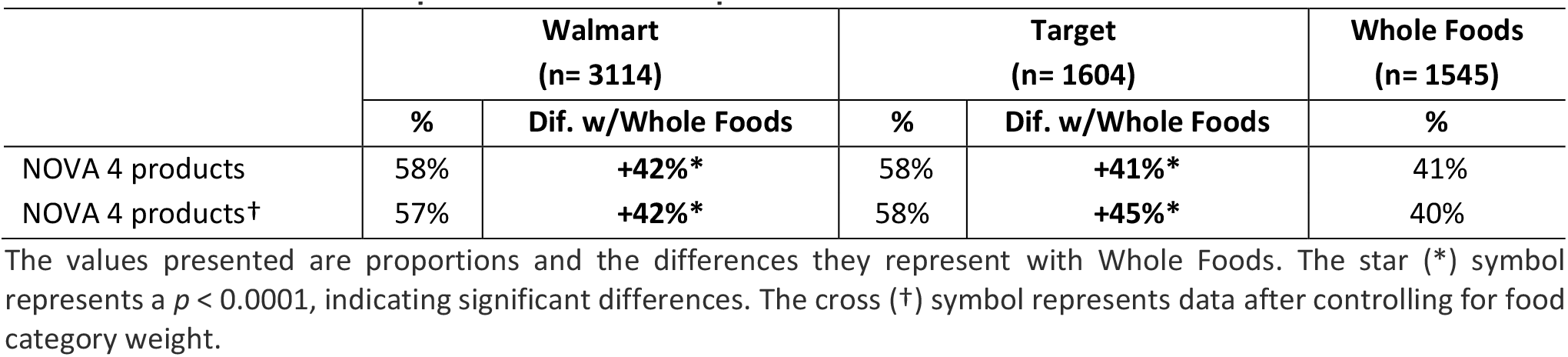
Prevalence of NOVA 4 products in the U.S. supermarkets.

|  | Walmart<br>(n= 3114) |  | Target<br>(n= 1604) |  | Whole Foods<br>(n= 1545) |
| --- | --- | --- | --- | --- | --- |
|  | % | Dif. w/Whole Foods | % | Dif. w/Whole Foods | % |
| NOVA 4 products | 58% | **+42%\*** | 58% | **+41%\*** | 41% |
| NOVA 4 products† | 57% | **+42%\*** | 58% | **+45%\*** | 40% |

The values presented are proportions and the differences they represent with Whole Foods. The star (\*) symbol represents a $p < 0.0001$ , indicating significant differences. The cross (†) symbol represents data after controlling for food category weight.

### 3.2 UPF markers in the United States

The average number of UPF markers found in the UPFs followed the same tendency. Walmart UPFs had the highest number of UPF markers, followed closely by Target and then trailed substantially by Whole Foods (Table 4). On average, we found 75% more UPF markers in the UPFs from Walmart vs. Whole Foods, and 57% more UPF markers in the foods from Target vs. Whole Foods.

**Table 4.**
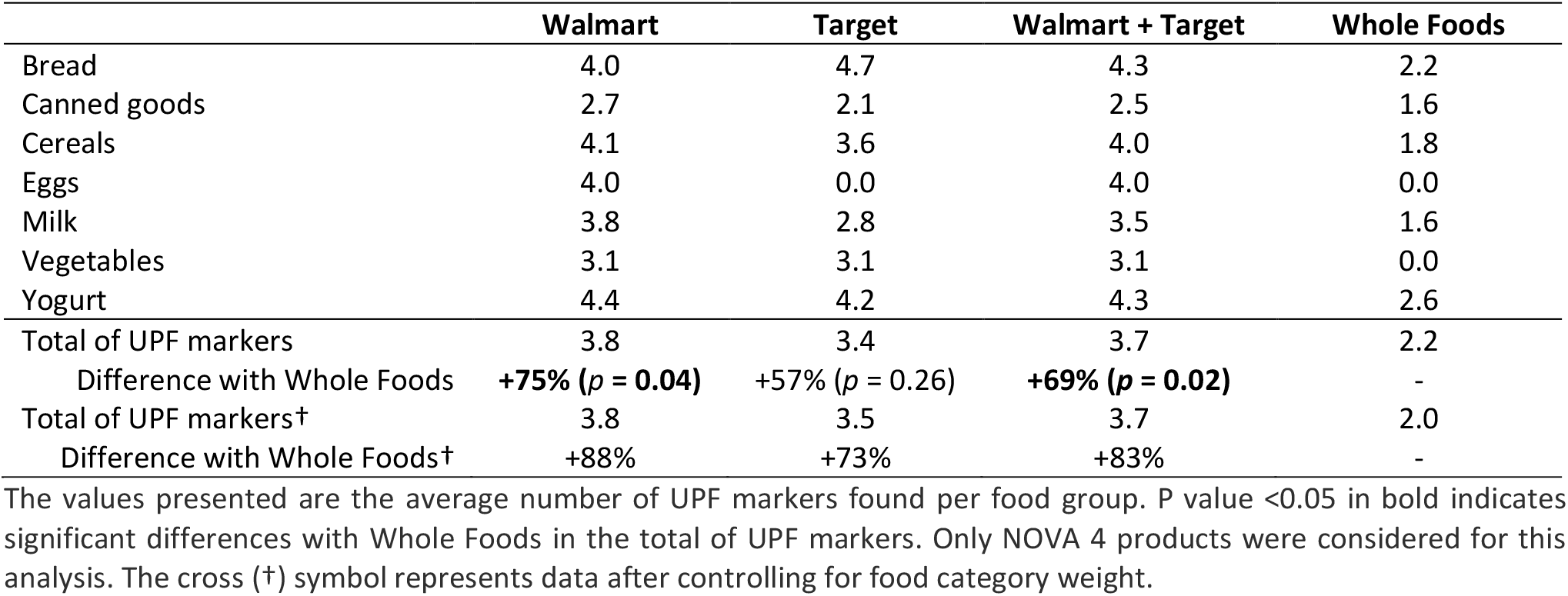
Number UPF markers found in the UPFs of each supermarket.

|  | Walmart | Target | Walmart + Target | Whole Foods |
| --- | --- | --- | --- | --- |
| Bread | 4.0 | 4.7 | 4.3 | 2.2 |
| Canned goods | 2.7 | 2.1 | 2.5 | 1.6 |
| Cereals | 4.1 | 3.6 | 4.0 | 1.8 |
| Eggs | 4.0 | 0.0 | 4.0 | 0.0 |
| Milk | 3.8 | 2.8 | 3.5 | 1.6 |
| Vegetables | 3.1 | 3.1 | 3.1 | 0.0 |
| Yogurt | 4.4 | 4.2 | 4.3 | 2.6 |
| Total of UPF markers | 3.8 | 3.4 | 3.7 | 2.2 |
| Difference with Whole Foods | **+75% ($p = 0.04$)** | +57% ($p = 0.26$) | **+69% ($p = 0.02$)** | - |
| Total of UPF markers† | 3.8 | 3.5 | 3.7 | 2.0 |
| Difference with Whole Foods† | **+88%** | **+73%** | **+83%** | - |

The values presented are the average number of UPF markers found per food group. P value $< 0.05$ in bold indicates significant differences with Whole Foods in the total of UPF markers. Only NOVA 4 products were considered for this analysis. The cross (†) symbol represents data after controlling for food category weight.

Specifically, 61% of Walmart UPFs have 3 or more UPF markers, compared with 56% in Target and 32% in Whole Foods (Table 5). There are 92% more UPF products with at least 3 markers at Walmart than at Whole Foods and 77% more at Target than Whole Foods.

**Table 5.** Amount of UPFs per number of UPF markers.

| Category | Walmart<br>(n= 1821) | Target<br>(n= 933) | Walmart + Target<br>(n= 2754) | Whole Foods<br>(n= 638) |
| --- | --- | --- | --- | --- |
| UPFs with 1 UPF marker | 386 (21%) | 240 (26%) | 626 (23%) | 283 (44%) |
| UPFs with 2 UPF markers | 320 (18%) | 169 (18%) | 489 (18%) | 152 (24%) |
| UPFs with 3 UPF markers | 267 (15%) | 135 (14%) | 402 (15%) | 85 (13%) |
| UPFs with ≥ 4 UPF markers | 848 (47%) | 389 (42%) | 1237 (45%) | 118 (18%) |
| UPFs ≥ 3 UPF markers | 1115 (61%) | 524 (56%) | 1639 (60%) | 203 (32%) |
| Difference with Whole Foods | **+92%\*** | **+77%\*** | **+88%\*** |  |

The values presented are the number of products per category and the proportion they represent over the total amount. The star (\*) symbol represents a $p < 0.0001$ indicating significant differences with Whole Foods in relation to the amount of UPFs with ≥3 UPF markers. Only NOVA 4 products were considered for this analysis.

### 3.3 UPF prevalence in mainstream supermarkets in the United States versus Europe

Based on the NOVA classification, 58% of staple products found at Walmart and Target combined (U.S.) were ultra-processed, while in Europe (France and Spain combined) this number was 41% (Table 6). Table 7 shows that this represented that the U.S. supermarkets have 41% more UPFs.

**Table 6.**
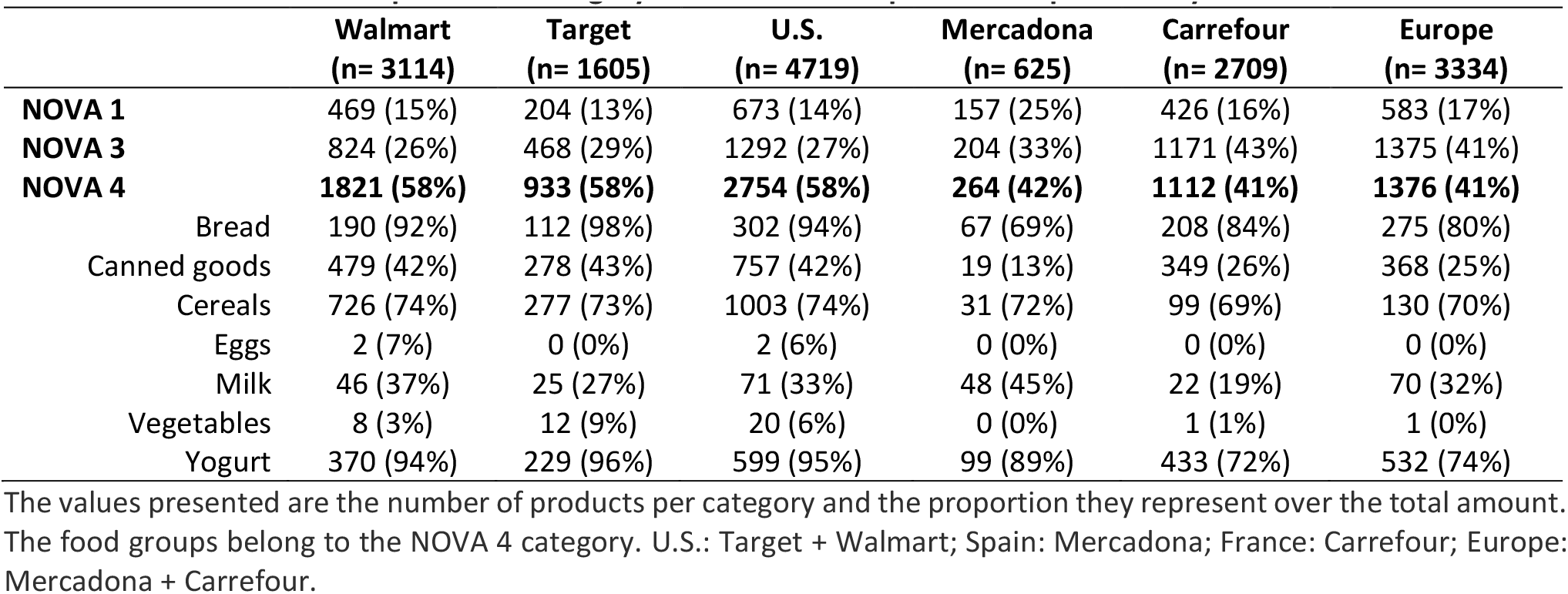
Product distribution per NOVA category in the selected supermarkets per country.

**Table 7.**
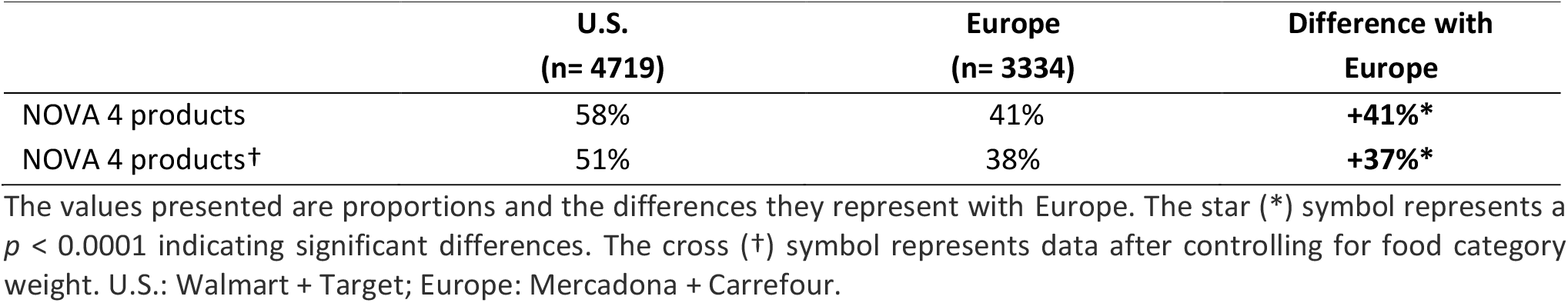
Prevalence of NOVA 4 products in the United States versus Europe.

|  | U.S.<br>(n= 4719) | Europe<br>(n= 3334) | Difference with<br>Europe |
| --- | --- | --- | --- |
| NOVA 4 products | 58% | 41% | **+41%\*** |
| NOVA 4 products† | 51% | 38% | **+37%\*** |

The values presented are proportions and the differences they represent with Europe. The star (\*) symbol represents a $p < 0.0001$ indicating significant differences. The cross (†) symbol represents data after controlling for food category weight. U.S.: Walmart + Target; Europe: Mercadona + Carrefour.

### 3.4 UPF markers in the mainstream supermarkets in the United States versus Europe

Once more, when analyzing the number of UPF markers we found the same tendency and the UPFs in the United States had 41% more UPF markers than in Europe (Table 8).

**Table 8.** Number of UPF markers found in the UPFs of each supermarket.

|  | Walmart | Target | U.S. | Mercadona | Carrefour | Europe |
| --- | --- | --- | --- | --- | --- | --- |
| Bread | 4.0 | 4.7 | 4.3 | 2.3 | 2.3 | 2.3 |
| Canned goods | 2.7 | 2.1 | 2.5 | 2.6 | 1.8 | 1.8 |
| Cereals | 4.1 | 3.6 | 4 | 2.2 | 3.0 | 2.8 |
| Eggs | 4.0 | - | 4.0 | - | - | - |
| Milk | 3.8 | 2.8 | 3.5 | 2.4 | 1.6 | 2.1 |
| Vegetables | 3.1 | 3.1 | 3.1 | - | 1.0 | 1 |
| Yogurt | 4.4 | 4.2 | 4.3 | 4.1 | 3.1 | 3.3 |
| Total of UPF markers | 3.8 | 3.4 | 3.65 | 3.0 | 2.5 | 2.6 |
| Difference with Europe |  |  | **+41% (p= 0.11)** |  |  |  |
| Total of UPF markers† | 3.8 | 3.5 | 3.7 | 2.9 | 2.5 | 2.6 |
| Difference with Europe† |  |  | **+44%** |  |  |  |

The values presented are the average number of UPF markers found per food group. P value <0.05 indicates significant differences with Europe in the total of UPF markers. Only NOVA 4 products were considered for this analysis. The cross (†) symbol represents data after controlling for food category weight.

Finally, Table 9 shows that up to 60% of UPF products in the United States have 3 or more UPF markers, whereas in Europe, this number decreases to 38%. This represented 57% more UPFs with over 3 UPF markers in the United States.

**Table 9.** Amount of UPFs per number of UPF markers.

|  | Walmart<br>(n= 1821) | Target<br>(n= 933) | U.S.<br>(n= 2754) | Mercadona<br>(n= 264) | Carrefour<br>(n= 1112) | Europe<br>(n= 1376) |
| --- | --- | --- | --- | --- | --- | --- |
| UPFs with 1 UPF marker | 386 (21%) | 240 (26%) | 626 (23%) | 78 (30%) | 440 (40%) | 518 (38%) |
| UPFs with 2 UPF markers | 320 (18%) | 169 (18%) | 489 (18%) | 53 (20%) | 282 (25%) | 335 (24%) |
| UPFs with 3 UPF markers | 267 (15%) | 135 (14%) | 402 (15%) | 47 (18%) | 173 (16%) | 220 (16%) |
| UPFs with ≥ 4 UPF markers | 848 (47%) | 389 (42%) | 1237 (45%) | 86 (32%) | 217 (19%) | 303 (22%) |
| UPFs ≥ 3 UPF markers | 1115 (61%) | 524 (56%) | 1639 (60%) | 133 (50%) | 309 (35%) | 523 (38%) |
| Difference with Europe |  |  | **+57%\*** |  |  |  |

The values presented are the number of products per category and the proportion they represent over the total amount. The star (\*) symbol represents a p <0.0001 indicating a significant difference with Europe in relation to the amount of UPFs with ≥3 UPF markers. Only NOVA 4 products were considered for this analysis.

## 4 DISCUSSION

Our research provides new evidence on the availability of food products contributing to the United States being the global leader in UPF consumption. While our findings highlight a pressing concern of public health in the United States, they also provide guidance to identify and implement corrective actions.

In line with the documented high UPF consumption in the United States (22), our research shows the high availability of UPFs among staple products (58%), especially in two of the largest retailers, Walmart, and Target (33), which showed 41% more UPFs than Whole Foods, known for being more expensive and focused on quality (27). High consumption levels may thus reflect the food stocking practices and availability in widely accessible supermarkets. This has been previously and similarly found in France revealing that UPF prevalence was higher in conventional stores than in organic stores (34).

Moreover, the prevalence of UPF markers also varies according to the type of supermarket in the United States (35), as confirmed by our results showing 69% more UPF markers in products found in budget-friendly supermarkets (Walmart and Target) compared to products found in premium supermarkets (Whole Foods). The number of UPF markers might be linked to the industry’s desire to save production costs (18) and produce ultra-palatable products, replacing unprocessed products in the shopping trolley. According to these findings and in line with other authors (36), considering the number of UPF markers in foods is a promising way to identify adverse products and empower consumers to easily identify UPFs as they shop. This could be useful to manufacturers, retailers, and the community and could set grounds for improving the health potential of highly influential businesses and food products.

According to our results, UPF markers reach even basic, staple products such as milk, bread, cereals, eggs, vegetables, yogurts, and canned goods, challenging U.S. citizens to eat healthfully in popular supermarkets even when avoiding items considered to be “junk” foods. Do consumers have to choose between affordable and healthy products? Our cross-country findings suggest not. U.S. main supermarkets have 41% more UPFs than the main supermarkets in France and Spain. Also, the number of UPF markers (cosmetic additives and other substances) is 41% higher than in Europe. Interestingly, the supermarket leaders we studied in Europe have the same UPF percentage as Whole Foods. Given that Mercadona and Carrefour have about the same market share as Walmart and Target, two of the biggest supermarkets in the United States (30,31) it seems that a tradeoff between health and affordability is not inevitable. In Europe, some of the largest retailers in each country have fewer UPFs and less UPF markers, compared with their U.S. counterparts.

UPFs are highly profitable for the food, beverage and restaurant industry sectors given the low-cost ingredients, long shelf-life, and powerful branding of these products (20). Moreover, despite the negative health outcomes (1), consumers continue choosing them, as UPFs are cheaper and more hyperpalatable than healthy options, among other reasons. Thus, the harmful cycle of product availability and consumer choice keeps going (4).

In light of our findings, expecting U.S. consumers to make healthy choices in an environment saturated with UPFs may be unrealistic. The current scenario is akin to asking individuals to make healthy lifestyle choices in an environment that promotes the opposite. The consumer, therefore, faces the dietary challenge of finding healthy and accessible options in popular supermarkets where UPFs prevail, even more so given the lack of clear definitions and awareness among consumers regarding what constitutes ultra-processed foods (37,38).

The current UPF situation in the United States compromises public health and calls for immediate action. The U.S. consumers’ choice is currently limited in subtle ways so that even basic, staple foods are likely to be ultra-processed. Given the significant impact of UPFs on public health and the challenges in altering consumer behavior in an environment dominated by these products, regulatory intervention seems necessary (39). History shows the effectiveness of such approaches (40). Such intervention could include stricter regulations on UPF content and labeling, public awareness campaigns about the health risks associated with UPFs, and incentives for supermarkets to promote healthier food options. While consumer education and empowerment are crucial (41), the scale of the problem requires a robust approach. Regulatory action, similar to that taken against tobacco in the public health initiatives in the 1960s and 70s (40), could significantly mitigate the health risks posed by UPFs and foster a healthier food environment. In addition, the success and extensive reach of big food retailers gives them huge potential as leaders for social impact in public health of the U.S. population.

Based on the general pricing trends of UPFs versus non-UPFs and the known price positioning of different supermarkets, our approach allows to address the economic dimension of consumer choices in the context of UPF prevalence without needing direct price comparisons for each product type. Variables, such as product price and consumer opinion, in association with health outcomes and UPF consumption and availability, would be interesting to explore.

## 5 CONCLUSION

UPFs in the United States dominate food supplies where least expected: in the staple foods for sale in major supermarkets. This situation is more severe in the country’s largest, mainstream retailers (Target and Walmart), and less problematic in a higher-end quality supermarket (Whole Foods). Our findings suggest that healthy food choices in the United States are compromised by the high availability and accessibility of UPFs, even among everyday products that constitute the dominant part of a population’s diet. Thus, U.S. citizens are challenged to choose between health and affordability. Moreover, the UPF prevalence in the United States is significantly higher than in Europe, as well as the number of UPF markers present in the UPFs. The European model shows that supermarkets can better balance health and affordability, with large-share supermarkets in Spain and France offering foods comparable to the higher-end U.S. stores. This suggests a potential path forward for U.S. supermarkets: revise product offerings to encourage healthier consumer choices. We believe that important steps toward this goal lie in government policies that prioritize less processed foods as well as consumer tools that empower them to identify UPFs and alternative, healthier products as they shop.

## Data Availability

All data produced in the present study are available upon reasonable request to the authors

## REFERENCES

1. Touvier M, da Costa Louzada ML, Mozaffarian D, Baker P, Juul F, Srour B. Ultra-processed foods and cardiometabolic health: public health policies to reduce consumption cannot wait. BMJ [Internet]. 2023 Oct 9 [cited 2024 Feb 1];e075294. Available from: https://www.bmj.com/content/383/bmj-2023-075294

2. Baraldi LG, Martinez Steele E, Canella DS, Monteiro CA. Consumption of ultra-processed foods and associated sociodemographic factors in the USA between 2007 and 2012: evidence from a nationally representative cross-sectional study. BMJ Open [Internet]. 2018 Mar 1 [cited 2024 Jan 5];8(3). Available from: https://pubmed.ncbi.nlm.nih.gov/29525772/

3. Mertens E, Colizzi C, Peñalvo JL. Ultra-processed food consumption in adults across Europe. Eur J Nutr [Internet]. 2022 Apr 3 [cited 2024 Feb 1];61(3):1521–39. Available from: https://pubmed.ncbi.nlm.nih.gov/34862518/

4. Monteiro CA, Cannon G, Levy RB, Moubarac JC, Louzada MLC, Rauber F, et al. Ultra-processed foods: what they are and how to identify them. Public Health Nutr [Internet]. 2019 Apr 1 [cited 2024 Jan 5];22(5):936–41. Available from: https://www.cambridge.org/core/journals/public-health-nutrition/article/ultraprocessed-foods-what-they-are-and-how-to-identify-them/E6D744D714B1FF09D5BCA3E74D53A185

5. Pagliai G, Dinu M, Madarena MP, Bonaccio M, Iacoviello L, Sofi F. Consumption of ultra-processed foods and health status: A systematic review and meta-Analysis [Internet]. Vol. 125, British Journal of Nutrition. Cambridge University Press; 2021 [cited 2024 Feb 1]. p. 308–18. Available from: https://pubmed.ncbi.nlm.nih.gov/32792031/

6. Davidou S, Christodoulou A, Frank K, Fardet A. A study of ultra-processing marker profiles in 22,028 packaged ultra-processed foods using the Siga classification. Journal of Food Composition and Analysis. 2021 Jun 1;99:103848.

7. Monteiro CA, Cannon G, Lawrence M, Laura Da Costa Louzada M, Machado PP. Ultra-processed foods, diet quality, and health using the NOVA classification system [Internet]. Food and Agriculture Organization of the United Nations (FAO). Rome; 2019 [cited 2024 Jan 28]. Available from: https://www.fao.org/3/ca5644en/ca5644en.pdf

8. Poti JM, Braga B, Qin B. Ultra-processed Food Intake and Obesity: What Really Matters for Health-Processing or Nutrient Content? [Internet]. Vol. 6, Current obesity reports. 2017 [cited 2024 Feb 1]. p. 420–31. Available from: https://pubmed.ncbi.nlm.nih.gov/29071481/

9. Delpino FM, Figueiredo LM, Bielemann RM, Da Silva BGC, Dos Santos FS, Mintem GC, et al. Ultra-processed food and risk of type 2 diabetes: A systematic review and meta-analysis of longitudinal studies [Internet]. Vol. 51, International Journal of Epidemiology. Oxford University Press; 2022 [cited 2024 Feb 1]. p. 1120–41. Available from: https://pubmed.ncbi.nlm.nih.gov/34904160/

10. Fiolet T, Srour B, Sellem L, Kesse-Guyot E, Allès B, Méjean C, et al. Consumption of ultra-processed foods and cancer risk: Results from NutriNet-Santé prospective cohort. BMJ (Online) [Internet]. 2018 [cited 2024 Feb 1];360. Available from: https://pubmed.ncbi.nlm.nih.gov/29444771/

11. Lane MM, Gamage E, Travica N, Dissanayaka T, Ashtree DN, Gauci S, et al. Ultra-Processed Food Consumption and Mental Health: A Systematic Review and Meta-Analysis of Observational Studies [Internet]. Vol. 14, Nutrients. MDPI; 2022 [cited 2024 Feb 1]. Available from: https://pubmed.ncbi.nlm.nih.gov/35807749/

12. Martínez Steele E, Popkin BM, Swinburn B, Monteiro CA. The share of ultra-processed foods and the overall nutritional quality of diets in the US: Evidence from a nationally representative cross-sectional study. Popul Health Metr [Internet]. 2017 Feb 14 [cited 2024 Feb 1];15(1). Available from: https://pubmed.ncbi.nlm.nih.gov/28193285/

13. Juul F, Simões BDS, Litvak J, Martinez-Steele E, Deierlein A, Vadiveloo M, et al. Processing level and diet quality of the US grocery cart: Is there an association? Public Health Nutr [Internet]. 2019 Sep 1 [cited 2024 Feb 1];22(13):2357–66. Available from: https://pubmed.ncbi.nlm.nih.gov/31190676/

14. WHO. World Health Organization. 2023 [cited 2024 Feb 2]. Noncommunicable diseases. Available from: https://www.who.int/news-room/fact-sheets/detail/noncommunicable-diseases#:~:text=Noncommunicable%20diseases%20(NCDs)%20kill%2041,%2D%20and%20middle%2Dincome%20countries.

15. CDC. Center for Disease Control and Prevention. 2023 [cited 2024 Feb 2]. Global Noncommunicable Diseases Fact Sheet | Division of Global Health Protection | Global Health | CDC. Available from: https://www.cdc.gov/globalhealth/healthprotection/resources/fact-sheets/global-ncd-fact-sheet.html

16. Lustig RH. Ultraprocessed Food: Addictive, Toxic, and Ready for Regulation. Nutrients [Internet]. 2020 Nov 5 [cited 2024 Feb 1];12(11):3401. Available from: https://www.ncbi.nlm.nih.gov/pmc/articles/PMC7694501/

17. Fardet A. Minimally processed foods are more satiating and less hyperglycemic than ultra-processed foods: a preliminary study with 98 ready-to-eat foods. Food Funct [Internet]. 2016 May 1 [cited 2024 Jan 29];7(5):2338–46. Available from: https://pubmed.ncbi.nlm.nih.gov/27125637/

18. Gupta S, Hawk T, Aggarwal A, Drewnowski A. Characterizing ultra-processed foods by energy density, nutrient density, and cost. Front Nutr [Internet]. 2019 May 28 [cited 2023 Dec 8];6:454858. Available from: https://pubmed.ncbi.nlm.nih.gov/31231655/

19. Cotter T, Kotov A, Wang S, Murukutla N. “Warning: ultra-processed” - A call for warnings on foods that aren’t really foods. BMJ Glob Health [Internet]. 2021 Dec 1 [cited 2024 Jan 5];6(12). Available from: https://pubmed.ncbi.nlm.nih.gov/34933866/

20. Monteiro CA, Cannon G, Lawrence M, Laura Da Costa Louzada M, Machado PP. Ultra-processed foods, diet quality, and health using the NOVA classification system Prepared by. Cad Saude Publica [Internet]. 2010 [cited 2024 Jan 5];26(11):2039–49. Available from: http://www.wipo.int/amc/en/mediation/rules

21. Enriquez JP, Gollub E. Snacking Consumption among Adults in the United States: A Scoping Review. Nutrients [Internet]. 2023 Apr 1 [cited 2024 Jan 5];15(7). Available from: https://www.ncbi.nlm.nih.gov/pmc/articles/PMC10097271/

22. Juul F, Parekh N, Martinez-Steele E, Monteiro CA, Chang VW. Ultra-processed food consumption among US adults from 2001 to 2018. Am J Clin Nutr [Internet]. 2022 Jan 1 [cited 2024 Jan 5];115(1):211–21. Available from: https://pubmed.ncbi.nlm.nih.gov/34647997/

23. Ozbun T. Statista. 2023 [cited 2024 Feb 2]. Walmart - Statistics & Facts | Statista. Available from: https://www.statista.com/topics/1451/walmart/

24. Leibtag E, Barker C, Dutko P. How Much Lower Are Prices at Discount Stores? United States Department of Agriculture [Internet]. 2010 Sep [cited 2024 Jan 5]; Available from: https://www.ers.usda.gov/webdocs/publications/44759/8076_err105.pdf?v=0

25. Walmart. Walmart History [Internet]. [cited 2023 Dec 8]. Available from: https://corporate.walmart.com/about/history

26. Smith P. Statista. 2023 [cited 2024 Jan 5]. Target - Statistics & Facts. Available from: https://www.statista.com/topics/1914/target/#topicOverview

27. Whole Foods. Our Values — Whole Foods Market UK [Internet]. [cited 2023 Dec 8]. Available from: https://www.wholefoodsmarket.co.uk/our-values

28. Shahbandeh M. Statista. 2023 [cited 2024 Feb 2]. Whole Foods Market - Statistics & Facts | Statista. Available from: https://www.statista.com/topics/2261/whole-foods-market/

29. Sarah Min. Which U.S. grocery chains offer the best value? - CBS News [Internet]. 2019 [cited 2024 Feb 9]. Available from: https://www.cbsnews.com/news/which-us-supermarkets-offer-the-best-value/

30. Statista Research Department. Statista. 2023 [cited 2024 Jan 5]. Leading supermarkets by market share France 2023. Available from: https://www.statista.com/statistics/535415/grocery-market-share-france/

31. Orús Abigail. Statista. 2023 [cited 2024 Jan 5]. Supermercados: cuota de mercado en España en 2022. Available from: https://es.statista.com/estadisticas/540894/porcentaje-de-ventas-de-los-grandes-supermercados-en-espana/

32. GoCoCo [Internet]. [cited 2024 Jan 29]. Available from: https://www.gococo.app/es/home

33. Ozbun T. Statista. 2023 [cited 2024 Jan 5]. U.S. supermarket & other grocery store sales 2022. Available from: https://www.statista.com/statistics/197626/annual-supermarket-and-other-grocery-store-sales-in-the-us-since-1992/

34. Davidou S, Frank K, Christodoulou A, Fardet A. Organic food retailing: to what extent are foods processed and do they contain markers of ultra-processing? Int J Food Sci Nutr [Internet]. 2022 [cited 2024 Feb 2];73(2):172–83. Available from: https://www.tandfonline.com/doi/abs/10.1080/09637486.2021.1966395

35. Ravandi B, Mehler P, Barabási AL, Menichetti G. GroceryDB: Prevalence of Processed Food in Grocery Stores. medRxiv [Internet]. 2022 Oct 12 [cited 2024 Jan 5]; Available from: https://www.medrxiv.org/content/10.1101/2022.04.23.22274217v2

36. Davidou S, Christodoulou A, Fardet A, Frank K. The holistico-reductionist Siga classification according to the degree of food processing: an evaluation of ultra-processed foods in French supermarkets. Food Funct [Internet]. 2020 Mar 26 [cited 2024 Feb 2];11(3):2026–39. Available from: https://pubs.rsc.org/en/content/articlehtml/2020/fo/c9fo02271f

37. Sarmiento-Santos J, Souza MBN, Araujo LS, Pion JMV, Carvalho RA, Vanin FM. Consumers’ Understanding of Ultra-Processed Foods. Foods [Internet]. 2022 May 1 [cited 2024 Jan 5];11(9). Available from: https://pubmed.ncbi.nlm.nih.gov/35564081/

38. Sadler CR, Grassby T, Hart K, Raats M, Sokolović M, Timotijevic L. Processed food classification: Conceptualisation and challenges. Trends Food Sci Technol [Internet]. 2021 Jun 1 [cited 2024 Jan 5];112:149–62. Available from: https://www.sciencedirect.com/science/article/pii/S0924224421001667

39. Zhang Y, Giovannucci EL. Ultra-processed foods and health: a comprehensive review. Crit Rev Food Sci Nutr [Internet]. 2023 [cited 2024 Jan 5];63(31). Available from: https://pubmed.ncbi.nlm.nih.gov/35658669/

40. Pollack Porter KM, Rutkow L, McGinty EE. The Importance of Policy Change for Addressing Public Health Problems. Public health reports [Internet]. 2018 Nov 1 [cited 2024 Jan 5];133(1). Available from: https://pubmed.ncbi.nlm.nih.gov/30426876/

41. Popkin BM, Barquera S, Corvalan C, Hofman KJ, Monteiro C, Ng SW, et al. Towards unified and impactful policies to reduce ultra-processed food consumption and promote healthier eating. The Lancet [Internet]. 2021 Jul 1 [cited 2024 Jan 5];9(7):462–70. Available from: https://pubmed.ncbi.nlm.nih.gov/33865500/

